# Predicting adult Attention Deficit Hyperactivity Disorder (ADHD) using vocal acoustic features

**DOI:** 10.1101/2021.03.18.21253108

**Authors:** Georg G. von Polier, Eike Ahlers, Julia Amunts, Jörg Langner, Kaustubh R. Patil, Simon B. Eickhoff, Florian Helmhold, Daina Langner

## Abstract

**Background:** It is a key concern in psychiatric research to investigate objective measures to support and ultimately improve diagnostic processes. Current gold standard diagnostic procedures for attention deficit hyperactivity disorder (ADHD) are mainly subjective and prone to bias. Objective measures such as neuropsychological measures and EEG markers show limited specificity. Recent studies point to alterations of voice and speech production to reflect psychiatric symptoms also related to ADHD. However, studies investigating voice in large clinical samples allowing for individual-level prediction of ADHD are lacking. The aim of this study was to explore a role of prosodic voice measures as objective marker of ADHD.

**Methods:** 1005 recordings were analyzed from 387 ADHD patients, 204 healthy controls, and 100 clinical (psychiatric) controls. All participants (age range 18-59 years, mean age 34.4) underwent an extensive diagnostic examination according to gold standard methods and provided speech samples (3 min in total) including free and given speech. Paralinguistic features were calculated, and random forest based classifications were performed using a 10-fold cross-validation with 100 repetitions controlling for age, sex, and education. Association of voice features and ADHD-symptom severity assessed in the clinical interview were analyzed using random forest regressions.

**Results and Conclusion:** ADHD was predicted with AUC = 0.76. The analysis of a non-comorbid sample of ADHD resulted in similar classification performance. Paralinguistic features were associated with ADHD-symptom severity as indicated by random forest regression. In female participants, particularly with age < 32 years, paralinguistic features showed the highest classification performance (AUC = 0.86).

Paralinguistic features based on derivatives of loudness and fundamental frequency seem to be promising candidates for further research into vocal acoustic biomarkers of ADHD. Given the relatively good performance in female participants independent of comorbidity, vocal measures may evolve as a clinically supportive option in the complex diagnostic process in this patient group.

## Introduction

Attention Deficit Hyperactivity Disorder (ADHD) is a neurodevelopmental condition defined by symptoms of inattention, hyperactivity, and impulsivity, which impair quality of life, social, and academic outcome [1, 2]. Core symptoms are often accompanied with emotional irritability, disorganized behavior and problems of self-concept, as well as in neuropsychological domains including executive functions and working memory. Comorbid psychiatric symptoms and disorders occur very frequently, especially in adults. The condition is highly prevalent worldwide with estimates of 5% prevalence in childhood and about 1.5% in adults resulting in functional and occupational impairments with sequelae like academic and social failure [3, 4]. Current ADHD diagnostic work up according to international guidelines [5, 6] is built around rater depended assessment tools (including a diagnostic interview as well as self- and third-party-reports and rating scales). These are however subjective procedures and therefore prone to biases [7]. Additionally, memory-recall biases occur when adults are asked to trace back childhood symptoms. Limitations of this current diagnostic gold standard have been published and the clinical practice is criticized for mis-, over- or under-diagnosing ADHD [8, 9], which can lead to the inappropriate prescription of medication, failure to prevent chronification, and poor psychosocial development [4]. Thus, a major aim in psychiatric research is to develop (objective) biomarkers, in order to help improve the diagnostic process and accuracy.

While various objective measurement techniques have provided valuable information in ADHD, none have yet been accepted into routine diagnostics, mostly because they lack convincing accuracy and/or have limited practicality as outlined in the following. Neuropsychological tests have detected impaired sustained attention and working memory deficits in patients with ADHD compared to healthy controls [10]. However, minimal differences between groups of patients and clinical controls have been reported given large intra-individual variation in performance [11]. Combined cognitive performance and actigraphy measures have shown reasonable correct classification rates for ADHD cases compared to healthy controls [12], however, separation from clinical controls is poor [13-15]. A huge body of research report EEG changes in ADHD, such as deflected theta/ beta ratio or an unstable sleep microstructure [16], but recent meta-analyses indicate a very limited diagnostic value of this approach [17, 18]. In neuroimaging, ADHD specific magnet resonance imaging (MRI) markers have been identified [19-21] and ADHD related changes in structural and functional connectivity have been reported [22-25]. Studies to investigate individual prediction have yielded lower accuracy in out of sample performance, partly due to variance related to different MRI scanners. Moreover, neuroimaging techniques are among the least practical to implement in routine care. In summary, while laboratory tests such as neuropsychological tests, EEG or MRI may add to the diagnostic process of ADHD in order to rule out somatic or metabolic differential diagnoses, a robust, clinically feasible ADHD biomarker has yet to be identified.

Recently, the analysis of voice and speech anomalies as a potential biomarker has gained considerable interest in neurologic [26] and psychiatric diseases [27-29]. The field recognizes the potential of an individual-level prediction using machine learning classification approaches and the possibility of objective, non-invasive, time and cost-efficient assessments in clinical settings including remote options (telephone or voice over the internet) [30]. Voice conveys information on multiple levels, such as volume, pitch, rate, and pauses. It reflects a broad underlying central nervous system involvement including networks and structures known to be affected in ADHD [31]. Neurophysiologically, speaking is shaped by vocal fold actions and sounds are filtered and articulated by sound production mechanisms of other articulators, as well as the pharynx, mouth, tongue and lips. Speaking is described as using more motor fibers than any other body activity, forming up to 30 phonetic segments per second [32], which underlies the intricate modulation and qualities of sound signals. Speech production is believed to involve extensive neural networks and be dependent on dopaminergic transmission, executive functions, and working memory [33]. Speech output is continuously being monitored by proprioceptive and auditory loops, [34-37] and deficits in speech production and auditory perception related to impairments in working memory and executive functions have been reported in ADHD (reviews at [34-37]).

Voice, perceived as a complex psychomotor task, should therefore naturally yield differences between ADHD and healthy controls. Some observations point into this direction: Children with ADHD and to some degree their parents poorly modulate voice volume, speak louder and for longer stretches, exhibiting signs of hyperfunctional voice disorder [38, 39]. They have higher subglottal pressure and lower transglottal airflow [38], which could result from heightened muscle tone in the glottis due to ADHD-associated increases in tonus [40, 41].

Taken together, in ADHD, voice is a rich source of information that may be used in individual diagnostic prediction in combination with machine learning methods, as has been studied in other psychiatric disorders such as schizophrenia [42] and depressive disorders [30] but so far not in ADHD. Given the (dopaminergic) alterations in brain functioning in ADHD, voice and speech production are possibly altered to a degree that may pertain to changes in speech production that could ultimately support clinical decision-making. The aim of this study is to determine whether prosodic voice features recorded in brief, clinically feasible speech tasks may allow for a differentiation of ADHD from healthy probands and possibly also probands with mental disorders other than ADHD using a machine learning based statistical approach.

## Methods

### Subjects

563 adult probands were recruited at our specialized adult ADHD outpatient clinic, 387 of which were subsequently diagnosed with ADHD (see table 1) and 100 of which were diagnosed with other mental disorders (see table S3 for diagnostic distribution in this group) but did not fulfill the diagnostic criteria for ADHD. 76 patients were excluded from further analyses due to positive drug screenings (n = 55), technical deficits of the recordings (n=7), or because they showed subclinical ADHD symptoms, but did not fulfill the criteria for ADHD or other mental disorders after the diagnostic evaluation (n = 14). All participants were asked to provide a voice recording before undergoing a standardized diagnostic procedure. In addition, 204 non-patient adults were recruited as healthy controls through public announcement. Healthy Controls had no history of past or present neuropsychiatric conditions. All participants were aged between 18 and 59 years and all gave written informed consent (see Box S1 in Supplement for full inclusion/ exclusion criteria). A detailed sample description is provided in Table 1, and S2. The authors assert that all procedures contributing to this work comply with the ethical standards of the relevant national and institutional committees on human experimentation and with the Helsinki Declaration of 1975, as revised in 2008. All procedures involving human subjects/patients were approved by the local ethics committee, the approval number is EA4/014/10. The study was registered as a clinical trial (ClinicalTrials.gov Identifier: NCT01104623).

**Table 1.** Sample description.

|  | male | female | total |
| --- | --- | --- | --- |
| All participants | 388 | 379 | 767 |
| Age (years, Mean; SD) | 34.3 (10.5) | 33.4 (9.8) | 34.4 (10.6) |
| ADHD (all) | 221 | 166 | 387 |
| ATT | 117 | 81 | 198 |
| COM | 104 | 85 | 189 |
| ADHD with comorbidity | 117 | 84 | 201 |
| HC | 75 | 129 | 204 |
| PC | 36 | 64 | 100 |
| Excluded | 56 | 20 | 76 |

ATT = inattentive ADHD subtype; COM = combined inattentive and hyperactive / impulsive ADHD subtype; HC = healthy controls; PC = psychiatric controls

### Diagnostic Procedure

ADHD diagnoses were obtained through a multi-informant, multi-method approach with psychological and medical assessment. All patients were assessed by trained and licensed psychologists and psychiatrists affiliated with the specialized ADHD adult outpatient clinic of the Charité University Hospital, Berlin, Germany. The **diagnostic procedure** was structured in accordance with recommended practice of national and international guidelines [43]. The diagnostic procedure represents a current “gold standard” for the diagnosis of ADHD in adults and included a clinical/diagnostic (DSM-) interview, review of client history (including developmental, medical, academic and social background) and relevant documentation, behavioral observations, completion of self- and third party-report and standardized ADHD-specific rating scales each for childhood and adult age. These evaluations were completed over two sessions with up to 8 hours of assessment (Table 2).

**Table 2.**
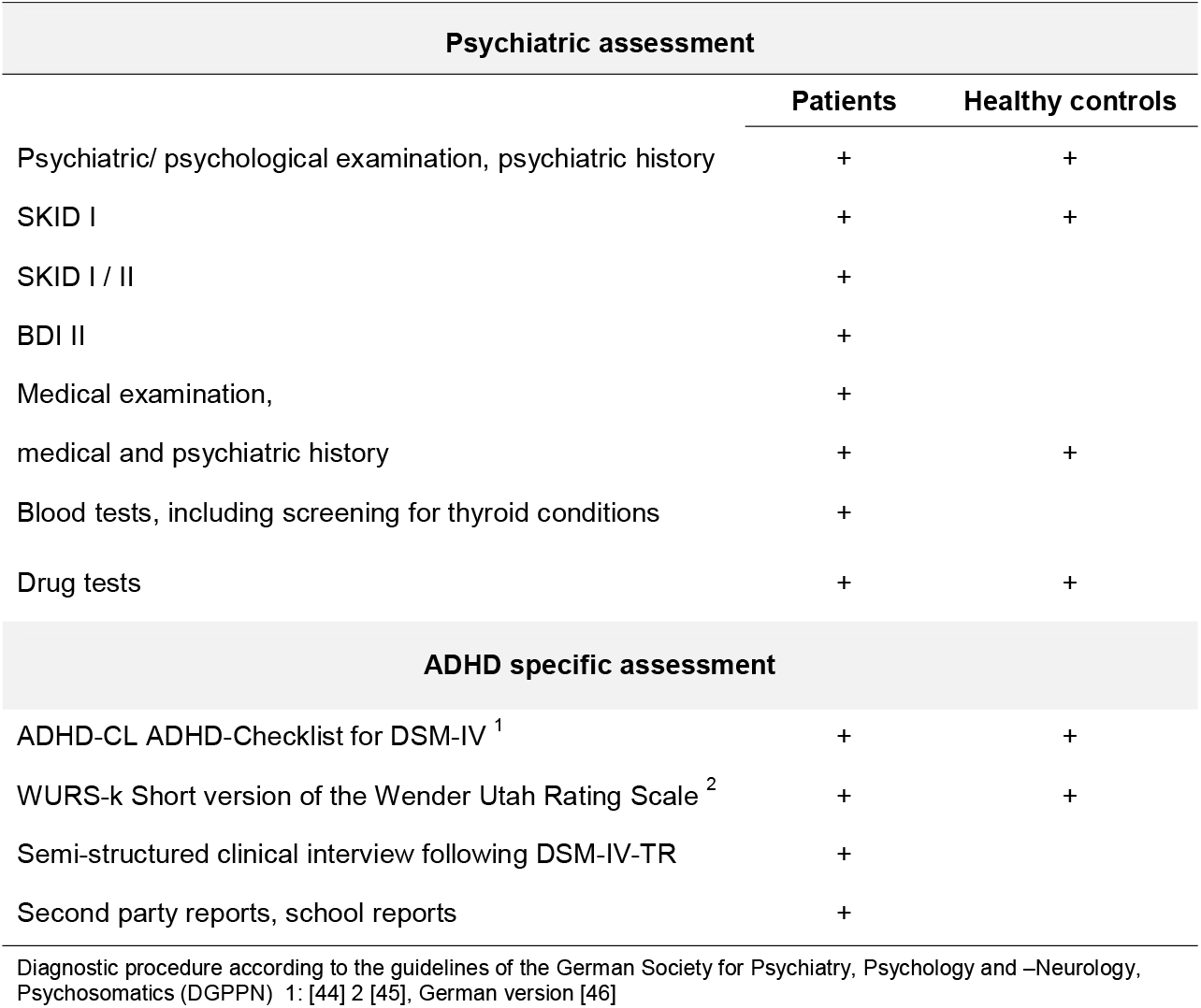
Diagnostic procedure.

| Psychiatric assessment |  |  |
| --- | --- | --- |
|  | Patients | Healthy controls |
| Psychiatric/ psychological examination, psychiatric history | + | + |
| SKID I | + | + |
| SKID I / II | + |  |
| BDI II | + |  |
| Medical examination, | + |  |
| medical and psychiatric history | + | + |
| Blood tests, including screening for thyroid conditions | + |  |
| Drug tests | + | + |
| ADHD specific assessment |  |  |
| ADHD-CL ADHD-Checklist for DSM-IV <sup>1</sup> | + | + |
| WURS-k Short version of the Wender Utah Rating Scale <sup>2</sup> | + | + |
| Semi-structured clinical interview following DSM-IV-TR | + |  |
| Second party reports, school reports | + |  |

Diagnostic procedure according to the guidelines of the German Society for Psychiatry, Psychology and –Neurology, Psychosomatics (DGPPN) 1: [44] 2 [45], German version [46]

To evaluate **ADHD-related symptom severity**, a clinical interview was performed by a senior clinician according to DSM-related criteria [43], the ADHD-DC scale [44]. In addition to 18 items covering inattention, hyperactivity, and impulsivity symptoms, the age of onset, symptoms related burden, general burden and reduced social contacts are rated on 0-3 point likert scales; 22 items in total, sum scores 0 - 66.

### Voice Recordings

High quality sound samples were recorded *prior* to the clinical evaluation so that participants and recording technicians were unaware of the diagnostic results. Since the probands had multiple clinical appointments and each appointment was preceded by the same recording task, multiple recordings of the same probands were recorded using the identical procedure. Participants spoke into a headset microphone (C555L AKG) and signals were recorded in 16-bit/22.5 kHz sampling rate on a PC followed by cutting and labeling procedures as preparation for the sound analysis via the “Fast Track” Interface (MediTECH Electronic GmbH). Prior to the actual recording, the instructions for the participants were recorded and played back through this interface in order to minimize the effects of interactions with the recording technicians and ensure a standardized recording procedure. All participants were asked to speak the following utterances: spoken single vowels and consonants, reading out of a given text, counting from one to ten (two trials) and free speech of around 120 seconds duration of a free topic, e.g. the last weekend/ holidays (see Table S4). All data were collected under identical recording conditions in a clinical examination office. From any given participant, a maximum of three recordings was included in the final analysis as separate data points.

From an initial 1029 voice recordings from 767 participants (including the participants that were later excluded), 24 recordings were excluded due to technical problems of the recordings (e.g., low volume) or due to multiple recordings, i.e., more than three recordings of the same person. From the remaining 1005 recordings, 85 recordings were excluded: (a) due to positive drug screenings of the probands (n = 60), because participants showed subclinical ADHD-symptoms but did not qualify for ADHD or other mental disorders (n = 24) or due to severe cold (n = 1). Thus, a total of 920 recordings were included in the classification steps, using 1 - 3 recordings of each participant.

### Feature generation and data analysis

Paralinguistic features were calculated, so only prosodic information but *no* semantic content was analyzed. Only the utterances based on free speech and counting were included in the analyses, spoken words and single sounds were not included into further analyses. Following a *Fourier* transformation of the voice recordings [47], paralinguistic features were calculated in two feature groups pertaining to the contour of loudness and the contour of fundamental frequency [48-50]. In brief, loudness was transformed to loudness as perceived by humans (i.e. Sone) based on the model of subjective loudness by Zwicker [51]. The loudness was then averaged over 24 different time frames with durations between 0.025 seconds and 4 seconds (see Box S2). Characteristics of the loudness curves like slopes, peaks, and curvatures of the respective time frames were calculated. For a second set of features, the fundamental frequencies were calculated and transformed on a logarithmic scale. The resulting curve was again averaged over the same 24 different time frames, and the characteristics (slopes, peaks, curvatures) of the melody curve belonging to the respective time frames were calculated [52]. To account for statistical effects related to the dimension time of the recordings, the curves of the respective features were fitted to multiple high-dimensional vectors resulting in a total of 6045 raw features.

Prior to the classification, initial filtering was performed to eliminate features with very low (< 2.2 e −16) variance (n=0) and variables with less than 100 unique values (n=4), resulting in 6041 features that were included in the further analyses. Random forest (RF) classifications were performed within MATLAB Version 2020a using the Tree-Bagger algorithm using 500 trees in 10-fold cross-validation repeated 100 times. Confounds (sex, age, education) were regressed from each feature separately using linear regression in a cross-validation consistent manner, i.e., the regression model was estimated on the training fold and applied to the training and validation folds and the residuals retained as confound-free features. Classification performance was estimated using the area under the receiver operating characteristic (AUC-ROC, in the following termed AUC).

To assess the prediction capacity of the voice features with regard to continuous variables such as ADHD symptom severity according to the ADHD-DC scale, age and education, we calculated random-forest based regression models with 100 trees also within the cross-validation folds described above with 100 repetitions. Mean absolute error of the regression and Pearson’s correlation between actual and predicted variables were calculated as performance measures. To compare Pearson’s coefficients, we first calculated Fisher r-to-z transformations and then performed a conservative version of t-tests as proposed by Nadeau and Bengio [53] to account for the possibility of bias related to cross-validation analyses.

In order to explore the prediction accuracy over different subgroups (i.e., gender, age-groups) thus further delineating the relevance of age and gender to voice analyses, we calculated group comparisons of male/ female subgroups and of two age groups of comparable sample size (median age 32 years). Group comparisons were performed using t-tests in a similar manner as described above. To evaluate the relevance of specific features and speech tasks for the classification of ADHD, the feature importance was assessed as defined by the permutation of out-of-bag predictions as implemented in MATLAB [54]. The top ten features were extracted for further evaluation.

## Results

### Descriptive results

As expected, ADHD probands showed a significantly higher symptom severity according to the sum score in the clinical interview than healthy and clinical control group probands (see Table 3). Comparisons with excluded subjects remained non-significant.

**Table 3.** Testpsychology.

|  | Clinical Interview<br>M ± SD | Inattention<br>M ± SD | Hyperactivity/<br>Impulsivity<br>M ± SD | W.U.R.S.-k<br>M ± SD | BDI - II<br>M ± SD |
| --- | --- | --- | --- | --- | --- |
| **ADHD, PC, HC** | 29.8 ± 16.6 | 4.9 ± 3.2 | 3.6 ± 2.9 | 30.1 ± 18.1 | 12.7 ± 9.1 |
| **ADHD** | 40.0 ± 8.3 | 7.0 ± 1.7 <sup>2</sup> | 4.9 ± 2.5 <sup>2</sup> | 38.9 ± 12.9 | 13.3 ± 8.9 |
| **ADHD - male** | 40.5 ± 8.3 | 7.0 ± 1.7 | 4.7 ± 2.5* | 39.4 ± 12.9 | 12.7 ± 8.7 |
| **ADHD - female** | 39.3 ± 8.3 | 6.9 ± 1.6 | 5.2 ± 2.6* | 38.3 ± 12.9 | 14.0 ± 9.2 |
| **ADHD - ATT** | 35.1 ± 7.3** | 7.0 ± 1.6 | 2.8 ± 1.6** | 36.6 ± 11.6** | 13.3 ± 9.0 |
| **ADHD - COM** | 44.7 ± 6.6** | 6.9 ± 1.8 | 7.0 ± 1.1** | 41.5 ± 13.7** | 13.0 ± 8.9 |
| **ADHD < 32 y <sup>1</sup>** | 40.2 ± 8.0 | 7.0 ± 1.5 | 4.7 ± 2.6 | 38.0 ± 11.8 | 13.7 ± 9.0 |
| **ADHD ≥ 32 y <sup>1</sup>** | 39.5 ± 8.9 | 6.8 ± 1.8 | 5.0 ± 2.5 | 39.8 ± 13.7 | 12.8 ± 8.8 |
| **HC** | 5.2 ± 4.4*** | 0.5 ± 0.8*** | 0.5 ± 0.8*** | 6.6 ± 6.5*** | 3.2 ± 2.8*** |
| **PC** | 23.3 ± 12.2 | 3.3 ± 2.5 | 2.7 ± 2.3 | 23.7 ± 13.8 | 14.9 ± 8.8 |
| **Excl. Particip.** | 35.9 ± 11.6 | 5.5 ± 2.4 | 4.5 ± 2.6 | 39.1 ± 15.4 | 16.5 ± 9.3 |

ATT = inattentive ADHD subtype; COM = combined inattentive and hyperactive / impulsive ADHD subtype; HC = healthy controls; PC = psychiatric controls; W.U.R.S.-k = Wender-Utah-Rating-Scale short form; BDI = Becks-Depression-Inventory; \* p < 0.05 in comparisons of sex; \*\* p < 0.001 in comparisons ATT vs COM; \*\*\* p < 0.001 in comparisons ADHD vs HC; 1: Comparisons between age-groups of ADHD-Participants remained non-significant; 2: Comparing the variance of Inattention and Hyperactivity with Levene's test indicates higher variance in Hyperactivity than in Inattention

Within the ADHD-group, the symptoms in the subscale “Inattention” were more pronounced than in the subscale “Hyperactivity/ Impusivity”, though of note, the latter showed a higher variance as indicated by a significant Levene’s test. Moreover, female ADHD-participants showed more pronounced “Hyperactivity” than male ADHD-participants, and this applied to participants from the ADHD-subgroup with combined symptoms (Inattention/ Hyperactivity-Impulsivity) compared to participants in the subgroup with predominantly “Inattention”. The comparisons of test-psychological results among age-groups of similar size revealed no significant group differences.

### Classification

The classification of patients and healthy controls using vocal per subject controlling for effects of age, sex and education resulted in a classification threshold invariant AUC = 0.76 (see Table 4, Figure 1).

**Table 4.** Results of the Classification ADHD vs HC.

| Group |  | AUC |
| --- | --- | --- |
| all |  | 0.76 |
| male |  | 0.69 |
| female |  | 0.82 |
| one recording/ participant |  | 0.76 |
| excl. stimulants |  | 0.76 |
| excl. comorbid ADHD |  | 0.75 |
| ADHD, subtype ATT |  | 0.78 |
| ADHD, subtype COM |  | 0.79 |
| **Age 18 – 31**<br>**(n = 385 recordings)** | all | 0.80 |
|  | male (n = 202) | 0.75 |
|  | Female (n = 183) | 0.86 |
| **Age 32 – 59**<br>**(n = 416 recordings)** | all | 0.71 |
|  | male (n = 193) | 0.59 |
|  | female (n = 223) | 0.74 |

ATT = inattentive ADHD subtype; COM = combined inattentive and hyperactive / impulsive ADHD subtype; HC = healthy controls

**Figure 1:**
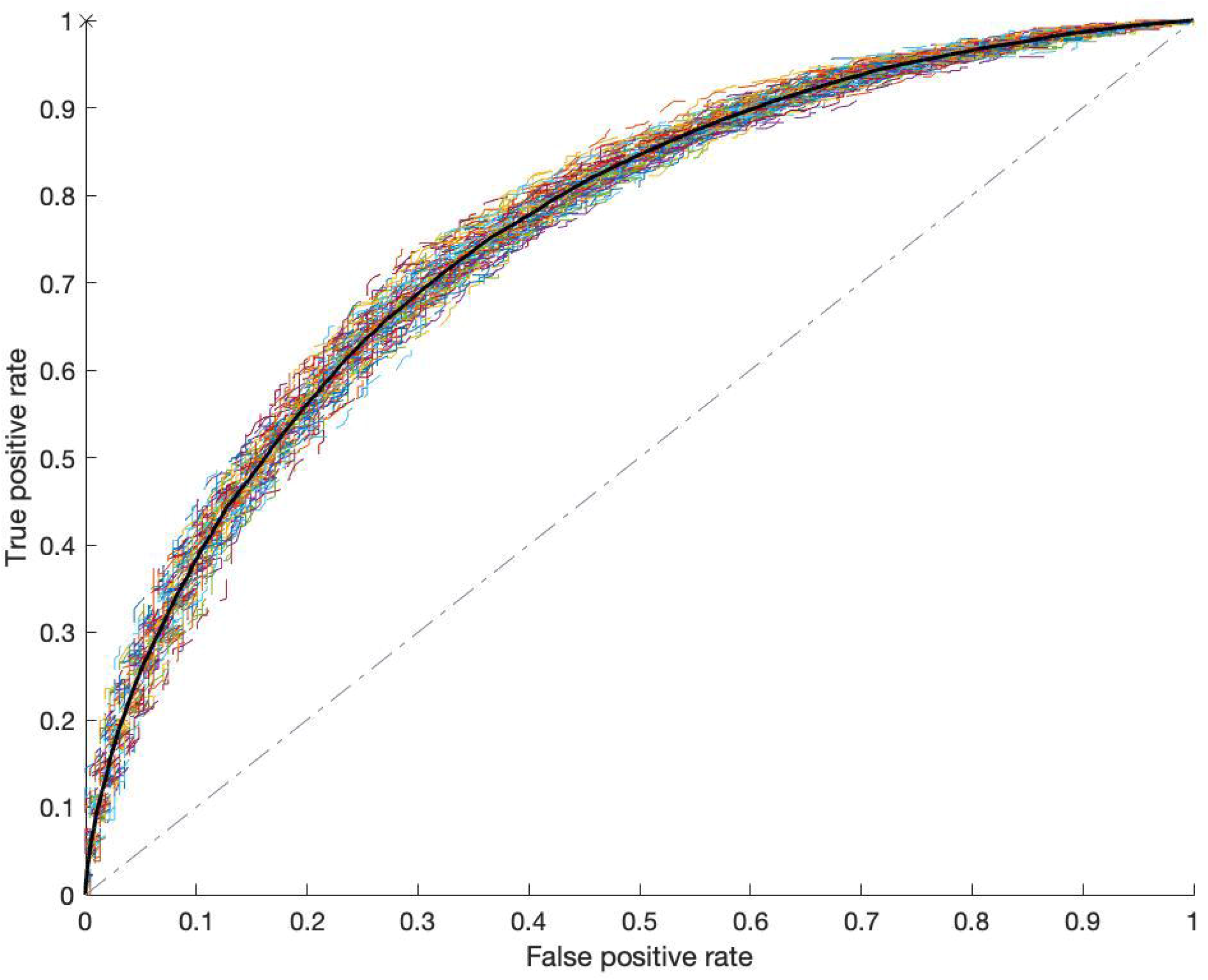
Classification ROCs for (all) ADHD patients versus healthy controls. Random-forest classification, 10-fold cross-validation, 100 replications; AUC = 0.76.

### ADHD-subtype, sex, age

The classification showed a largely similar performance in subjects with combined symptoms (i.e., including hyperactivity and impulsivity) as compared to predominantly inattentive subjects. Analyzing both sexes separately pointed toward a higher AUC in female than in male participants (*t* = 3.3, *p* = 0.001). A superior performance in female probands was also evident when analyzing separate age groups (age < 32: *t*_*male/ female*_ = 2.2, *p* = 0.03; age > 32: *t*_*male/ female*_ = 2.3, *p* = 0.03, see also Table 4) and in the non-comorbid ADHD sample (male: AUC = 0.68; female AUC= 0.79; *t* = 3.3, *p* = 0.001).

The comparisons of age groups (median age 32 years) indicated a superior classification performance in the younger sample (AUC = 0.80 vs AUC = 0.71; *t* = 2.0, *p* = 0.04), with the highest performance in young female participants (AUC = 0.86).

### Comorbidity, clinical controls, number of speech samples, medication

The classification of a subsample of ADHD subjects with no comorbidity resulted in a comparable AUC-ROC of 0.75, indicating that comorbidities do not affect the prediction considerably.

Calculating the classification of ADHD participants against a psychiatric control group, that presented for the evaluation of ADHD, but did not fulfill the clinical criteria of ADHD, showed a much lower prediction accuracy (AUC = 0.59). Of note, the psychiatric control group still showed elevated symptoms of ADHD as indicated by much higher mean scores in the clinical interview of ADHD compared to HC (mean sum score psychiatric controls: 23 vs 5 in HC; mean sum score 40 in ADHD, see table 3).

Notably, the performance using a single recording per subject was identical compared to including multiple recordings per subject. Excluding subjects taking ADHD medication (methylphenidate, atomoxetine) before the voice recording likewise did not change the classification performance, although the number of affected recordings was modest (n = 71; 8.9% of the analyzed sample).

### Correlations of symptom severity using random forest regression

The results are given in Table 5. The correlations of true and predicted symptoms performed in the *ADHD-group only* indicated associations of voice features and ADHD symptom severity *r* = 0.27, *t* = 6.7, *p* < 0.001), that seemed to be related to hyperactive/ impulsive symptoms to a larger extent than to inattention (*r* _hyper_ = 0.36; *r* _inattention_ = 0.16; comparison: *t* = 3.6, *p* <0.001). However, hyperactivity showed a larger variance than inattention.

**Table 5.** Correlation coefficients of true and predicted clinical variables.

|  | Group | n | *r* | *t* | MAE |
| --- | --- | --- | --- | --- | --- |
| ADHD Clinical Interview | all | 876 | 0.45 | 14.7 | 12.5 |
| ADHD Clinical Interview | female | 457 | 0.49 | 12.1 | 12.9 |
| ADHD Clinical Interview | male | 419 | 0.29 | 6.3 | 12.0 |
| - Hyperactivity | all | 915 | 0.36 | 11.6 | 2.3 |
| - Inattention | all | 915 | 0.39 | 12.9 | 2.6 |
| Education | all | 920 | 0.33 | 10.5 | 0.7 |
| Age | all | 920 | 0.52 | 18.5 | 7.2 |
| BDI | all | 701 | 0.24 | 6.6 | 7.1 |
| ADHD Clinical Interview | adhd | 554 | 0.27 | 6.7 | 6.3 |
| - Hyperactivity | adhd | 571 | 0.36 | 9.2 | 2.0 |
| - Inattention | adhd | 571 | 0.16 | 3.9 | 1.3 |

Pearsons's correlation coefficients are given; MAE = Mean absolute error; BDI = Becks Depression Inventory; $p$ -value for all analyses $< 0.001$

Extending the analysis of ADHD symptom severity and voice features to *all participants* (ADHD, PC, HC), the correlation of true and predicted symptom score was higher (*r* = 0.45, *t* = 14.7, *p* < 0.001), and this was particularly the case in female probands (*r*_female_ = 0.49; r_male_= 0.29; comparison *t* = 3.4, *p* <0.001). Moreover, no difference in correlations between true and predicted symptom scores of inattention/ hyperactive/ impulsive symptoms occurred.

### Confounding variables

To analyze confounders that likely play a role in voice analysis, we performed predictions of sex, age and education (= highest educational level) using RF-classification and RF-regression as appropriate in the whole sample (ADHD, PC, HC). Sex was classified with AUC = 0.98. The correlations of true and predicted age (*r* = 0.52, *t* = 18.5, *p* < 0.001) and education (*r* = 0.33, *t* = 10.5, *p* < 0.001) likewise pointed to relevant associations with voice features and were therefore included in all analyses as confounders of no interest. Moreover, sex (*t* = 5.4, *p* < 0.001), age (*t* = −1.7, *p* = 0.09) and education (*t* = 5.9, *p* < 0.001) were associated with ADHD-status and were therefore included as covariates of no interest as described above.

As depression has been linked to changes of loudness in past research [30, 55], we calculated associations of depression scores with voice features (*r* = 0.24, *t*=6.6, *p* < 0.001). However, depression scores were not related to ADHD symptom severity, thus we did not include it as covariate in the main analysis. Including depression severity as covariate in a secondary analysis did not change the results.

### Analysis of feature importance for the prediction of ADHD

The ten most important features were equally (50%) distributed to free speech (2 minutes) based on loudness changes over time frames between 0.020 – 0.252 seconds and changes of fundamental frequency in the task “counting one to ten” in timeframes of 0.02 to 0.1 seconds.

## Discussion

In the context of a growing interest in analyzing voice and speech as potential biomarkers of mental health [56-59], to our knowledge this is the first study to investigate the possible value of paralinguistic features in a large, heterogeneous sample of adults with ADHD using a machine learning based statistical approach. Our results support and extend previous findings of paralinguistic abnormalities in ADHD and point toward the possibility of predicting ADHD in unseen individuals, in particular in female probands in early adulthood.

### General performance in the context of the current literature

Our classifier showed a good prediction accuracy to differentiate ADHD from HC, reflected by an overall AUC = 0.76 and this is also reflected in regression analyses with associations of true and predicted symptom severity in the whole sample and within the ADHD group respectively. Since this is the first study to predict adult ADHD using paralinguistic features, in the following, we discuss the results in the context of (a) the current literature on the prediction of psychiatric diseases from voice data and (b) approaches to predict ADHD from biological signals other than voice.

Previous studies in the emerging field of automatic assessments of mental disorders using speech were restricted to mood and psychotic disorders [59] and showed comparable prediction accuracies as reported in this study: Depression can be predicted from (paralinguistic) voice features with an accuracy of about 80% [60]. The prediction of schizophrenia has been successful using linguistic features such as semantic relatedness of individuals with schizophrenia or those at a high risk to develop acute symptoms [42, 61-64]. Looking at the studies that have investigated the machine learning based prediction of ADHD from *other* biological signals points towards a similar performance of approaches based on neuropsychological performance [65], EEG-measures [55, 66], questionnaires [67] or resting state fMRI [68-70] as compared to our findings. In summary, the findings in this study are broadly comparable to previous research using voice to predict mental disorders or other biological signals to predict ADHD.

### Differentiation from clinical controls and comorbidity

Moreover, correlations between true and predicted symptom severity scores, both in the whole sample (ADHD, PC, HC; *r* = 0.45) and within the ADHD group alone (*r* = 0.27), indicate that voice features were able to predict ADHD-symptom *severity*. The continuous association of voice-features and ADHD-severity however may help to understand the lower prediction accuracy when calculating the classification of ADHD against clinical controls: the clinical control group presented in the outpatient clinic for a diagnostic workup to rule out ADHD and did show markedly elevated ADHD symptoms as compared to healthy controls. However, after an extensive diagnostic procedure, the participants in this group were not diagnosed with ADHD but with various other mental disorders, mostly affective disorders. Thus, since the classifier seems to be sensitive to lower levels of ADHD-symptoms as indicated by the RF-regression analyses of symptom severity above, the classification is likely less suited to differentiate between ADHD-suspect subjects with lower symptoms and subjects that fulfil the criteria of ADHD. Moreover, the clinical control group was heterogenous with regards to mental disorders and this may add to the limited differentiation from ADHD. Future studies with larger sample size may be able to predict ADHD with higher precision and possibly be able to better differentiate ADHD participants from participants with low or moderate ADHD symptoms. Likewise, other studies investigating potential biomarkers of ADHD point to a restricted differentiation from clinical controls such as studies of neurological tests in combination with actigraphy [71-74], meta-analyses evaluating fMRI [75] and resting-state fMRI studies to predict ADHD respectively [76, 77].

A strength of the current approach may be related to a robust classification performance of ADHD in the presence of comorbid mental disorders: the classifier showed a similar performance to differentiate ADHD from healthy controls *with and without* comorbidity. Regarding the role of comorbidity, we additionally controlled the analyses for depression symptom severity with no changes in the AUC. Depression is a frequent lifetime comorbidity in ADHD [78] and is known to influence paralinguistic features, such as reduced pitch range, speaking rate and intonation [30]. Of note, participants with severe clinical comorbidity, such as schizophrenia or severe affective disorders were excluded from the analysis. Overall, the limited influence of comorbidity in this study is encouraging in terms of possible clinical applications, replication of our results pending: adult ADHD patients often present with latent or lifetime comorbid conditions [79], thus compromising the diagnostic accuracy of ADHD in clinical practice [80]. Since our data point to a relatively robust accuracy particularly in younger female participants - possibly independent of comorbidity - automated voice analysis could evolve as a valuable addition to support the diagnostic processes, considering marked diagnostic challenges amongst others due to comorbidity in this patient group [81].

### Inattention and Hyperactivity/ Impulsivity

Evaluating the prediction accuracy of inattentive and hyperactivity/ impulsivity symptom scores, the RF-regression analyses pointed to a higher correlation of true and predicted symptoms of hyperactivity than of inattention in the ADHD group. Importantly, we noted a higher variance of the hyperactivity/ impulsivity symptoms than of inattention in the ADHD group, as indicated by a significant Levene’s test. Thus, the higher correlation of true and predicted hyperactivity/ impulsivity scores may to some extent be explained by a better ability for the RF regression to learn from a sample with larger variance. Also, using the whole sample (i.e., ADHD, PC, HC), inattention and hyperactivity subscales showed similar prediction scores of true and predicted symptoms (with similar variance of both variables in this group). Moreover, the higher correlation of true and predicted hyperactivity/ impulsivity symptoms did not translate into a significantly better prediction of the subgroup with combined (inattention and hyperactive/ impulsive) symptoms.

Recent research has emphasized the study of underlying traits of ADHD such as cognitive traits, e.g., executive functioning and temperament/personality features as an avenue to a create more informative phenotypes [82]. Studies to investigate cognitive traits such as executive functions and speech production [83] have found that high executive functioning performance relates to faster and more accurate word retrieval in adults and more accurate articulatory output children [60]. Articulation requires complex motor execution, that is dependent on self-regulation and inhibitory control. Since articulation is related to loudness and fundamental frequency [84], these features could in part be related to difference in executive functions between ADHD and healthy controls.

With regard to temperament/ personality traits as a second important symptom cluster of ADHD, prior research similarly has pointed to associations of personality traits and prosody of speech [30, 42], including associations of impulsivity and fundamental frequency/ jitter [85], that are related to the features in this study. Nilsen *et al*. [39] reported higher speech volumes and pitch to be related to decreased inhibitory control, a key component of impulsivity. Moreover, a higher motor activity in ADHD, may result into higher subglottal pressures that have been reported in children with ADHD [38]. In summary, a relation of prosodic speech abnormalities and ADHD traits such as deficits in executive functioning or hyperactivity/ impulsivity seems plausible and should be evaluated in more detail in future research.

### Sex and age

Sex was classified using the voice features with a high accuracy (AUC = 0.98), similar to other recent investigations using prosodic voice features and different classification approaches [86-88]. Furthermore, our data indicate a better classification performance of ADHD in female over male probands, and this pertained to subsamples with different age-groups and non-comorbid ADHD-probands. To some extent the sex related differences may be explained by slightly higher hyperactivity scores in female ADHD, but other measures of symptom severity or comorbidity did not differ between sexes. Thus, we assume that the voice features vary between male and female probands and are possibly more pronounced in female ADHD probands, though this has to be investigated in future studies including the underlying biological mechanisms. Other studies on voice in psychiatric disorders have mostly not differentiated between sex due to limited sample sizes. However, investigating sex separately has been recommended [59].

With respect to the role of age in the prediction performance, the features in this study were able to predict age with a comparable if slightly lower performance to other published approaches to investigate the prediction of age using prosodic features [89, 90]. Research on changes of prosody in aging indicates higher scores of hoarseness, instability and breathiness in higher age [91, 92], reflected by a diminished harmony to noise ratio [93]. Moreover, with increasing age, the subglottal pressure decreases in line with a decrease in overall muscle mass, and proband compensate this with increased expiratory airflow [94]. In ADHD, some studies report increased hoarseness [38, 95] and one study reports increased subglottal pressure and decreased airflow in children [38]. Thus, healthy probands in higher age may show symptoms such as hoarseness, a trait that may help differentiate ADHD from healthy controls. Moreover, ADHD probands in higher age may show decreased subglottal pressure, possibly resulting in a smaller difference from healthy controls. These hypotheses state a possible framework for the interpretation of the decreased classification performance in higher age. Since the ADHD-related findings have only been studies in children so far, our hypotheses should be tested in future research.

### Stimulants

Stimulants have been reported to impact voice and prosody in few small studies, including reports of lower fundamental frequency and increased jitter [96, 97], and increased hoarseness [98], In our data, we did not note differences with regard to classification accuracy, however the subsample with intake of stimulants was relatively small.

### Features and speaking tasks

Our results of changes in loudness support the limited number of previous studies that identified voice and speech anomalies in ADHD, especially pertaining to loudness changes in ADHD, as reviewed in 2016 [34]. Breznitz *et al*. reported differences in temporal speech patterns and physical features of vocalization in boys with ADHD [35] compared to clinical (reading disabilities) and healthy controls. Increased loudness, hoarseness and breathiness were identified in ADHD children compared to healthy controls [36, 37]. In particular, children with combined ADHD type were louder, showed lower fundamental frequency, had more straining voices, as well as more hoarseness and breathiness while speaking, compared to healthy controls. Due to a higher subglottal pressure and lower transglottal airflow in ADHD, abuse and misuse of the voice count as risk factors for functional dysphonias [38]. We assume that even subtle changes in voice based on higher subglottal pressure in ADHD patients may results in loudness features to play an important role in the classification of ADHD vs healthy probands.

Moreover, our data indicated that speech test selection plays an important role in the classification. Five of the top ten features were related to counting (from one to ten). While it is known that characteristics of paralinguistic features differ in different languages [99], the type of speech task also influences vocal features. Cognitive function, recently shown to differ in ADHD as discussed above [60], is involved in speech production depending on the complexity of speech tasks. Moreover, vocal variability differs between spontaneous speech and reading aloud in patients with depression [100]. Further, the complexity of spontaneous speech tasks plays an important role; Cohen *et al*. [66] observed greater vocal variability during a picture description task compared with one recalling autobiographical memories. Thus, a combination of free speech and speaking tasks may play an important role for the classification of ADHD and should be included in future trials.

## Conclusion

In summary, we were able to differentiate ADHD patients from healthy controls using voice features with good results in particular in younger and female probands. Strengths of the present study were the large, heterogeneous sample, inclusion of healthy and psychiatric controls and application of gold standard diagnostic procedures. We detected ADHD associated vocal patterns that probably reflect a disorder related vocal hyperfunction. These may be related to individual personality traits such as impulsivity and impaired executive functioning in ADHD. Replication pending, a particular strength of the voice-based approach might be the independence of psychiatric comorbidity, that very frequently occurs in ADHD and complicates the diagnostic process. In combination with a high feasibility and low cost to record and analyze voice, we see a high potential value for clinical practice especially in female and in younger probands and strongly encourage further investigation. A larger study sample including a larger psychiatric control group would be needed to achieve a better differentiation from clinical controls with low or moderate ADHD symptoms. However, the clinical value might be more to help the clinician to include potential differential diagnoses than excluding differential diagnoses based on one test. Moreover, in future research, the classification performance may increase with the inclusion of additional distinct speech features, such as speech pauses and the utilization of additional linguistic measures such as verbal fluency. Taken together, we consider voice analyses a promising avenue to support the diagnostic process in adult ADHD.

## Data Availability

The data that support the findings of this study are available from PeakProfiling GmbH with certain restrictions. Restrictions apply to the availability of these data, which were used under licence for this study. Please contact co-author JL with requests. The code of the classification is available from the corresponding author (GP) upon reasonable request.

## Acknowledgements

The Study was funded by the German Federal Ministry of Economics and Technology, ZIM program (www.zim-bmwi.de), as a collaboration between Charite and MediTech Electronics GmbH (www.meditech.de). The calculations were performed with computing resources granted by RWTH Aachen University under project lect0046. This study was supported by the European Union’s Horizon 2020 Research and Innovation Program under Grant Agreement No. 945539 (HBP SGA3) and No. 826421 (Virtual Brain Cloud). The authors wish to thank Michael Colla and Laura Gentschow, who enabled and led the study together with DL. We thank Paula Kunze and Inga Leerhoff for their great support in data collection and study coordination. DL wishes to thank Rolf Dietmar Wolf, Gerlafingen, Switzerland, for the inspiration and support in the very first steps to design the study. We thank all participants for their willingness and efforts to participate.

## Disclosures

EA participated and received payments in the national advisory board ADHD of Shire/Takeda. JL is co-founder and CTO of PeakProfiling GmbH. He created audio-features used in this study, that are intellectual property of PeakProfiling GmbH. FH received payments by PeakProfiling GmbH.

## Notes

### Clinical Trial

NCT01104623

### Author Declarations

The authors assert that all procedures contributing to this work comply with the ethical standards of the relevant national and institutional committees on human experimentation and with the Helsinki Declaration of 1975, as revised in 2008. All procedures involving human subjects/patients were approved by the ethics committee of the Charite Universitatsmedizin Berlin, Berlin, Germany, the approval number is EA4/014/10.

