## Supplemental Information for "Predicting adult Attention Deficit Hyperactivity Disorder (ADHD) using vocal acoustic features"

### Supplementary Information

**Data storage:** Data are stored at Charité Universitaetsmedizin Berlin, Department of Psychiatry, Campus Benjamin Franklin, Berlin, Germany.

**Box S1** Inclusion and Exclusion criteria

**Inclusion Criteria:**

1. Age 18-59.
2. Written informed consent.
3. Adult ADHD / Non-ADHD/ other disorders according to the fourth version of the Diagnostic and Statistical Manual for Mental Disorders (DSM-IV)
4. Normal vocal functioning, negative history for voice or laryngeal disorders.
5. Currently euthyroid condition; in case of past history of thyroid disease and need for current pharmacological treatment, patient needs to be on stable medication for at least 4 weeks.

**Exclusion Criteria:**

1. Severe mental conditions such as schizophrenia and psychotic disorders not otherwise specified, severe depressive episode, severe substance use disorder.
2. Positive drug tests.
3. Severe medical condition such as epilepsy.

**Box S2** Length of feature timewindows in ms:

20, 25, 32, 40, 50, 63, 79, 100, 126, 159, 200, 252, 318, 400, 504, 635, 800, 1008, 1270, 1600, 2016, 2540, 3200, 4032

**Table S1** Distribution of recordings

| | **male** | **female** | **total** |
|---|---|---|---|
| All recordings | 505 | 501 | 1005 |
| ATT | 167 | 123 | 290 |
| COM | 149 | 134 | 283 |
| ADHD with comorbidity | 175 | 129 | 304 |
| ADHD with stimulants | 32 | 39 | 71 |
| Healthy Controls | 79 | 149 | 228 |
| Psychiatric Controls | 43 | 76 | 119 |
| Excluded | 66 | 19 | 85 |

ATT = inattentive ADHD subgroup; COM = combined inattentive and hyperactive / impulsive ADHD subgroup; HC = healthy controls

**Table S2** Education

| | No school certificate | High School Certificate | High school advanced certificate (German Abitur) | Further education/ university |
|---|---|---|---|---|
| All participants | 10 | 268 | 257 | 225 |
| ATT | 1 | 76 | 66 | 55 |
| COM | 6 | 81 | 65 | 37 |
| HC | 0 | 38 | 72 | 94 |
| PC | 1 | 41 | 31 | 27 |
| Excluded | 2 | 32 | 23 | 12 |

ATT = inattentive ADHD subtype; COM = combined inattentive and hyperactive / impulsive ADHD subtype; HC = healthy controls; PC = psychiatric controls

**Table S3** Diagnostic distribution of clinical controls

| **Diagnosis** | **n** |
|---|---|
| Addiction disorder (e.g. games) | 13 |
| Schizophrenia | 3 |
| Bipolar disorder | 1 |
| Depressive episode | 49 |
| Dysthymia | 12 |
| OCD, PTSD (ICD-10 Chapter 4) | 35 |
| Specific phobia | 16 |
| Social phobia | 11 |
| Anorexia nervosa, Bulimia nervosa | 3 |
| Borderline PD | 9 |
| Narcissistic PD | 13 |
| Avoidant-restrictive PD | 10 |
| other PD | 17 |

OCD = Obsessive compulsive disorder; PTSD = post-traumatic stress disorder; PD = Personality disorder

**Table S4** Recorded utterances from each participant

| **Utterance** | **mean duration (s)** |
|---|---|
| Free speech | 120 |
| Counting 1-10, two trials | 22 |
| Reading out single words | 20 |
| Sounds (3 sec) a: i: u: ç f s n | 21 |
| Recorded material per participant | 183 |
